# The Effects of “Fangcang, Huoshenshan, and Leishenshan” Hospitals and Temperature on the Mortality of COVID-19

**DOI:** 10.1101/2020.02.26.20028472

**Authors:** Yuwen Cai, Tianlun Huang, Xin Liu, Gaosi Xu

## Abstract

**Background:** In December 2019, a novel coronavirus disease (COVID-19) broke out in Wuhan, China, however, the factors affecting the mortality remain unclear.

**Methods:** Thirty-two days of data that were shared by China National Health Commission and China Weather Net were collected using standard forms. The difference in the mortality of confirmed and severe cases before and after the use of “Fangcang, Huoshenshan, and Leishenshan” makeshift hospitals (MSHs) was tested using Mann-Whitney U test. We also studied whether air temperature (AT) could affect the above outcomes of COVID-19 cases by performing Spearman’s analysis.

**Results:** Eight days after the use of MSHs, the mortality of confirmed cases was significantly decreased both in Wuhan (*U* = 1, *P* < 0.001) and Hubei (*U* = 0, *P* < 0.001), while in non-Hubei regions, as a contrast, the mortality of confirmed cases remained unchanged (*U* = 40, *P* = 0.139). However, another eight days later, changes in the mortality in non-Hubei regions also became significant (*U* = 73, *P* = 0.039). Mortality of confirmed cases was found to be significantly correlated with AT both in Wuhan (*r* = −0.441, *P* = 0.012) and Hubei (*r* = −0.440, *P* = 0.012).

**Conclusions:** Our findings indicated that both the use of MSHs and the rise of AT were beneficial to the survival of COVID-19 cases.

## Introduction

In early December 2019, a novel coronavirus disease (COVID-19, previously known as 2019-nCoV) induced by severe acute respiratory syndrome coronavirus 2 (SARS-CoV-2) broke out in Wuhan, China.^1 2^ This newly discovered coronavirus had been confirmed to have human-to-human transmissibility^3^ and has now spread across the country.^4^ However, the mortality caused by COVID-19 had been reported to be unbalanced in different regions.^4^ Briefly speaking, the mortality in Wuhan city was generally higher than that in other cities, and the mortality in Hubei Province was generally higher than that in other provinces. The specific reason needed to be investigated so that we can better contain the epidemic.

Wuhan as the source of the epidemic in China, was under great pressure of treatment despite the assistance received from all over the country. Many patients in Wuhan were unable to see a doctor, could not be hospitalized previously. The medical resources consumed by rescuing such patients would then further compress the treatment space of other patients. Such a vicious circle induced by inappropriate resource allocation might be one of the reasons for the high mortality in Wuhan. In addition, by reviewing the outbreak of severe acute respiratory syndrome (SARS) in Guangdong in 2003, we could find that the SARS epidemic gradually subsided with the warming of the weather, and was basically controlled until the warm April and May. Therefore, we assume that different air temperature (AT) in different regions might also contributed to the unbalanced mortality.

Up to February 5, 2020, the first three Fangcang, Huoshenshan, and Leishenshan makeshift hospitals (MSHs) had been put into use in Wuhan.^5^ The Fangcang hospitals, which belong to field mobile medical systems, are composed of a number of movable cabins. They have multiple functions such as emergency treatment, surgical disposal, clinical examination and so on. In case of any public health emergency, the cabins can rush to the scene as soon as possible, and then in situ expand to a class II hospital.^6^ In the present study, we aim to research whether these MSHs could reduce the mortality induced by COVID-19. In addition, we also studied whether the AT could improve the survival of COVID-19 cases.

## Methods

### Data collection and mortality calculation

From January 21 to February 21, the number of each day’s total cases confirmed by nucleic acid test, severe cases and new deaths in Wuhan city, Hubei Province and non-Hubei regions (as a contrast so as to reduce bias) were collected by two authors independently. All the above data were available on the official website of National Health Commission of the people’s Republic of China (http://www.nhc.gov.cn/). Growth rate of confirmed cases was calculated by dividing the new confirmed cases by the total confirmed cases on the previous day. Each day’s mortality was calculated by dividing the number of new deaths by the average number of confirmed cases on that day and the previous day, and mortality of severe patients was calculated by the number of severe cases. Each day’s AT were collected from China Weather Net (http://www.weather.com.cn/), and the AT of Hubei Province was represented by the average AT of its cities.

### Statistical analysis

The data of each region was divided into group A (after the hospitals were built) and group B (before the hospitals were built). Since the sample size was small (less than 50), the normality of data was determined using Shapiro–Wilk test, and *P* value > 0.05 was considered as normally distributed.^7^ If the outcomes of two groups were both normally distributed, Student’s t test would be performed to compare the difference between groups, and if the outcomes of at least one group was skewed distribution, Mann-Whitney U test would be performed instead.^8^ We compared data of eight days, sixteen days after the use of MSHs, respectively, with the data of sixteen days before the use of MSHs. As for the correlation analysis, if the AT and the corresponding outcome were both normally distributed, Pearson correlation analysis would be performed to investigate their correlation, otherwise, Spearman’s correlation analysis would be performed instead.^9^ SPSS 26.0 statistical software (IBM, New York, USA) was used for statistical data processing, and GraphPad Prism 8.3 (GraphPad Software Inc., New York, USA) was applied for plotting graphs. All tests were two-sided, and *P* value < 0.05 was considered as significant.

## Results

### Mortality difference before and after the use of MSHs

Each day’s total confirmed cases, severe cases, new deaths and AT in different regions were summarized in **Table 1**. As shown in **Supplementary Table S1**, for each outcome, at least one group was skew distribution (*P* < 0.05), so Mann-Whitney U test was performed to compare the difference between groups. The results of eight days after the use of MSHs are shown in **Figure 1** and **Table 2**. The growth rate of confirmed cases was significantly decreased both in Wuhan (*U* = 27, *P* = 0.023) and Hubei (*U* = 23, *P* = 0.012), but in non-Hubei regions, as a contrast, changes were also significant (*U* = 3, *P* < 0.001). The mortality of confirmed cases was significantly decreased both in Wuhan (*U* = 1, *P* < 0.001) and Hubei (*U* = 0, *P* < 0.001), while in non-Hubei regions, as a contrast, the mortality of confirmed cases remained unchanged (*U* = 40, *P* = 0.139). Similarly, the mortality of severe cases was significantly decreased in Hubei (*U* = 0, *P* < 0.001), while in non-Hubei regions, changes were not significant (*U* = 33, *P* = 0.056).

**Table 1:** Each day’s total confirmed cases, severe cases, new deaths and AT in different regions

| Date | Wuhan |  |  | Hubei |  |  |  | Non-Hubei regions |  |  |
| --- | --- | --- | --- | --- | --- | --- | --- | --- | --- | --- |
|  | Total confirm-<br>ed cases | Daily<br>deaths | AT | Total confirm-<br>ed cases | Total severe<br>cases | Daily<br>deaths | AT | Total confirm-<br>ed cases | Total severe<br>cases | Daily<br>deaths |
| 20-Jan | 258 | 6 |  | 270 | 51 | 6 |  | 21 | 17 | 0 |
| 21-Jan | 363 | 3 | 6.0 | 375 | 65 | 3 | 5.5 | 65 | 37 | 0 |
| 22-Jan | 425 | 8 | 4.0 | 444 | 71 | 8 | 4.7 | 127 | 24 | 0 |
| 23-Jan | 495 | 6 | 5.0 | 549 | 129 | 7 | 4.9 | 281 | 48 | 1 |
| 24-Jan | 572 | 15 | 5.5 | 729 | 157 | 15 | 4.5 | 558 | 80 | 1 |
| 25-Jan | 618 | 7 | 3.0 | 1052 | 192 | 13 | 3.3 | 923 | 132 | 2 |
| 26-Jan | 698 | 18 | 2.0 | 1423 | 290 | 24 | 2.3 | 1321 | 171 | 0 |
| 27-Jan | 1590 | 22 | 2.5 | 2567 | 690 | 24 | 2.4 | 1948 | 286 | 2 |
| 28-Jan | 1905 | 19 | 3.5 | 3349 | 899 | 25 | 3.9 | 2625 | 340 | 1 |
| 29-Jan | 2261 | 25 | 5.5 | 4334 | 988 | 37 | 5.6 | 3377 | 382 | 1 |
| 30-Jan | 2639 | 30 | 6.0 | 5486 | 1094 | 42 | 6.5 | 4206 | 433 | 1 |
| 31-Jan | 3215 | 33 | 6.5 | 6738 | 1294 | 45 | 7.2 | 5053 | 501 | 1 |
| 1-Feb | 4109 | 32 | 8.5 | 8565 | 1562 | 45 | 7.5 | 5815 | 548 | 0 |
| 2-Feb | 5142 | 41 | 8.5 | 9618 | 1701 | 56 | 7.4 | 7587 | 595 | 1 |
| 3-Feb | 6384 | 48 | 6.0 | 10990 | 2143 | 64 | 6.3 | 9448 | 645 | 0 |
| 4-Feb | 8351 | 49 | 7.0 | 12627 | 2520 | 65 | 7.7 | 11697 | 699 | 0 |
| 5-Feb | 10117 | 52 | 9.0 | 14314 | 3084 | 70 | 8.3 | 11988 | 775 | 3 |
| 6-Feb | 11618 | 64 | 5.0 | 15804 | 4002 | 69 | 3.8 | 13181 | 819 | 4 |
| 7-Feb | 13603 | 67 | 4.5 | 19835 | 5195 | 81 | 4.2 | 11939 | 906 | 5 |
| 8-Feb | 14982 | 63 | 5.5 | 20993 | 5247 | 81 | 6.4 | 12745 | 941 | 8 |
| 9-Feb | 16902 | 73 | 7.0 | 22160 | 5505 | 91 | 7.7 | 13822 | 979 | 6 |
| 10-Feb | 18454 | 67 | 7.5 | 25087 | 6344 | 103 | 8.2 | 12539 | 989 | 5 |
| 11-Feb | 19558 | 72 | 9.0 | 26121 | 7241 | 94 | 9.2 | 12679 | 963 | 3 |
| 12-Feb | 30043 | 82 | 11.0 | 43455 | 7084 | 107 | 10.7 | 9071 | 946 | 12 |
| 13-Feb | 32959 | 88 | 13.0 | 46806 | 9278 | 108 | 12.6 | 8942 | 926 | 5 |
| 14-Feb | 34289 | 77 | 11.0 | 48175 | 10152 | 105 | 10.2 | 8698 | 901 | 4 |
| 15-Feb | 35314 | 110 | 0.5 | 49030 | 10396 | 139 | 0.5 | 8386 | 876 | 3 |
| 16-Feb | 36385 | 76 | 2.0 | 49847 | 9797 | 100 | 3.8 | 8087 | 847 | 5 |
| 17-Feb | 37152 | 72 | 5.0 | 50338 | 10970 | 93 | 6.3 | 7678 | 771 | 5 |
| 18-Feb | 38020 | 116 | 7.5 | 50633 | 11246 | 132 | 7.3 | 7172 | 731 | 4 |
| 19-Feb | 37994 | 88 | 8.0 | 49665 | 11178 | 108 | 8.2 | 6638 | 686 | 6 |
| 20-Feb | 37448 | 99 | 10.0 | 48730 | 10997 | 115 | 9.9 | 6235 | 636 | 3 |
| 21-Feb | 36680 | 90 | 9.0 | 47647 | 10892 | 106 | 9.2 | 5637 | 585 | 3 |

AT, air temperature

**Table 2:** The difference before and after the use of MSHs

|  | Group | Mann-Whitney U test |  |  |
| --- | --- | --- | --- | --- |
|  |  | Median (QU, QL) | *U* value<sup>a</sup> | *P* value<sup>b</sup> |
| GRW | Before | 0.205 (0.165, 0.271) |  |  |
|  | After 8 days | 0.115 (0.093, 0.165) | 27 | = 0.023 |
|  | After 16 days | 0.050 (0.022, 0.121) | 27 | < 0.001 |
| GRH | Before | 0.268 (0.158, 0.346) |  |  |
|  | After 8 days | 0.091 (0.056, 0.224) | 23 | = 0.011 |
|  | After 16 days | 0.035 (0.007, 0.097) | 23 | < 0.001 |
| GRNH | Before | 0.326 (0.240, 0.879) |  |  |
|  | After 8 days | -0.002 (-0.094, 0.080) | 3 | < 0.001 |
|  | After 16 days | -0.043 (-0.088, 0.005) | 3 | < 0.001 |
| MCW (%) | Before | 1.152 (0.877, 1.768) |  |  |
|  | After 8 days | 0.410 (0.343, 0.513) | 1 | < 0.001 |
|  | After 16 days | 0.312 (0.234, 0.425) | 1 | < 0.001 |
| MCH (%) | Before | 0.893 (0.617, 1.447) |  |  |
|  | After 8 days | 0.409 (0.322, 0.450) | 0 | < 0.001 |
|  | After 16 days | 0.274 (0.220, 0.416) | 0 | < 0.001 |
| MCNH (%) | Before | 0.023 (0.001, 0.103) |  |  |
|  | After 8 days | 0.042 (0.033, 0.062) | 40 | = 0.153 |
|  | After 16 days | 0.049 (0.038, 0.063) | 73 | = 0.039 |
| MSH (%) | Before | 3.978 (3.196, 7.337) |  |  |
|  | After 8 days | 1.622 (1.411, 1.756) | 0 | < 0.001 |
|  | After 16 days | 1.337 (1.002, 1.657) | 0 | < 0.001 |
| MSNH (%) | Before | 0.230 (0.001, 0.758) |  |  |
|  | After 8 days | 0.557 (0.503, 0.806) | 33 | = 0.061 |
|  | After 16 days | 0.533 (0.463, 0.623) | 66 | = 0.019 |

<sup>a</sup> *U* value was used to test the significance of the difference
<sup>b</sup> *P* < 0.05 was considered as significantly different
Abbreviations: QL, lower quartile; QU, upper quartile; GRW, growth rate of confirmed cases in Wuhan; After 8 days, after the use of MSHs for 8 days; After 16 days, after the use of MSHs for 16 days; Before: before the use of MSHs; MCW: mortality of confirmed cases in Wuhan; GRH: growth rate of confirmed cases in Hubei; MCH: mortality of confirmed cases in Hubei; MSH: mortality of severe cases in Hubei; GRNH: growth rate of confirmed cases in non-Hubei regions; MCNH: mortality of confirmed cases in non-Hubei regions; MSNH: mortality of severe cases in non-Hubei region

**Figure 1:**
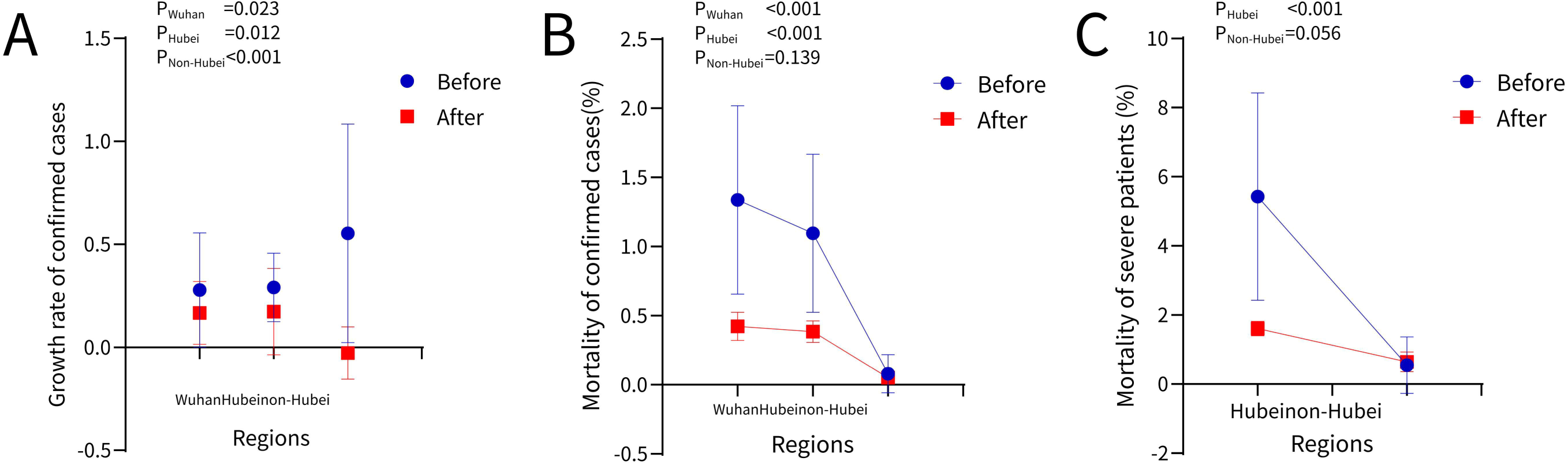
Comparisons between eight days after the use of MSHs and sixteen days before the use of MSHs. (**A**): comparisons of the growth rate of confirmed cases in Wuhan, Hubei, and non-Hubei regions. (**B**): comparisons of the mortality of confirmed cases in Wuhan, Hubei, and non-Hubei regions. (**C**): comparisons of the mortality of severe patients in Wuhan, Hubei, and non-Hubei regions. Before: before the use of MSHs. After: after the use of the use of MSHs.

The results of sixteen days after the use of MSHs were shown in **Figure 2** and **Table 2**. Results of the growth rate of confirmed cases was basically consistent with that of the first eight days. Changes in the mortality of confirmed cases were significant both in Wuhan (*U* = 1, *P* < 0.001) and Hubei (*U* = 0, *P* < 0.001), but not like the results of the first eight days, changes in non-Hubei regions became significant (*U* = 73, *P* = 0.038). Similarly, changes in the mortality of severe cases in Hubei were significant (*U* = 0, *P* < 0.001), while in non-Hubei regions, changes also became significant (*U* = 66, *P* = 0.019) this time.

**Figure 2:**
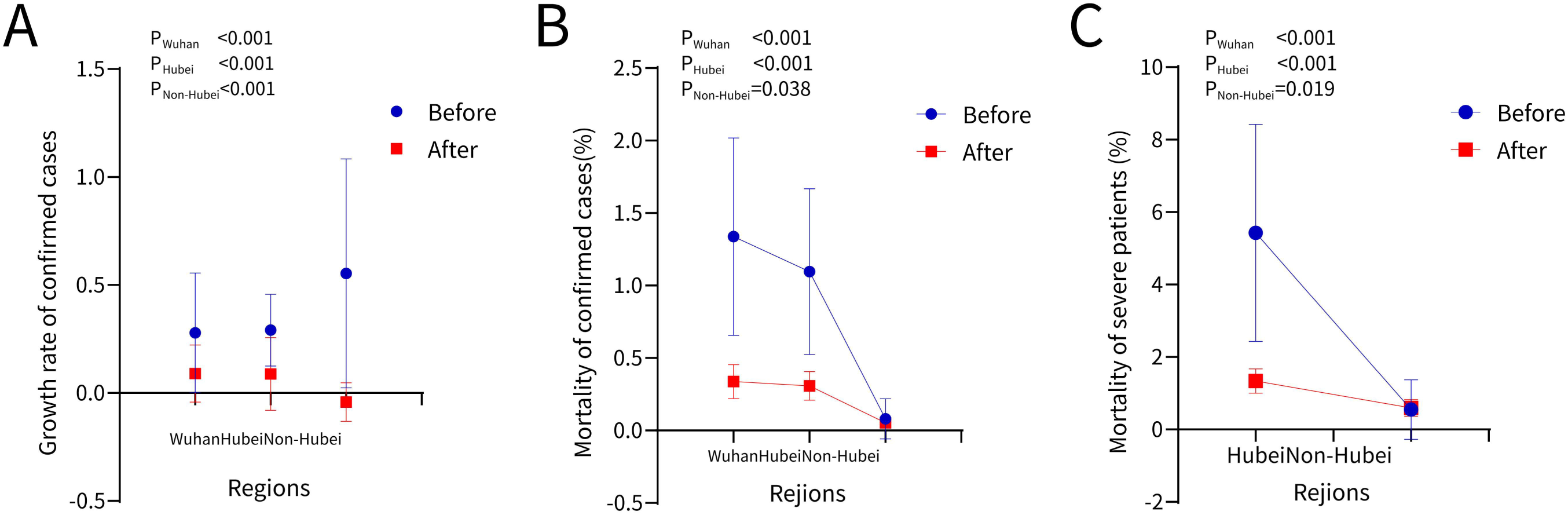
Comparisons between sixteen days after the use of MSHs and sixteen days before the use of MSHs. (**A**): comparisons of the growth rate of confirmed cases in Wuhan, Hubei, and non-Hubei regions. (**B**): comparisons of the mortality of confirmed cases in Wuhan, Hubei, and non-Hubei regions. (**C**): comparisons of the mortality of severe patients in Wuhan, Hubei, and non-Hubei regions. Before: before the use of MSHs. After: after the use of the use of MSHs.

### Correlation between AT and outcomes

Except AT, all outcomes were skew distribution as shown in **Supplementary Table S2**. Therefore, Spearman’s analysis was used to investigate their correlation among AT, and the results are shown in **Figure 3**. For the growth rate of confirmed cases, it was not significantly related to AT no matter in Wuhan (*P* = 0.730) or in Hubei (*P* = 0.062). But for the mortality of confirmed cases, its correlation between AT was considered to be significant both in Wuhan (*r* = −0.441, *P* = 0.012) and Hubei (*r* = −0.440, *P* = 0.012). The mortality of severe patients was also found to be significantly related to AT (*r* = −0.421, *P* = 0.016). That is to say, if the AT rises 1 Celsius, the mortality of confirmed cases would decrease 0.44% and the mortality of severe cases would decrease 0.42% on average.

**Figure 3:**
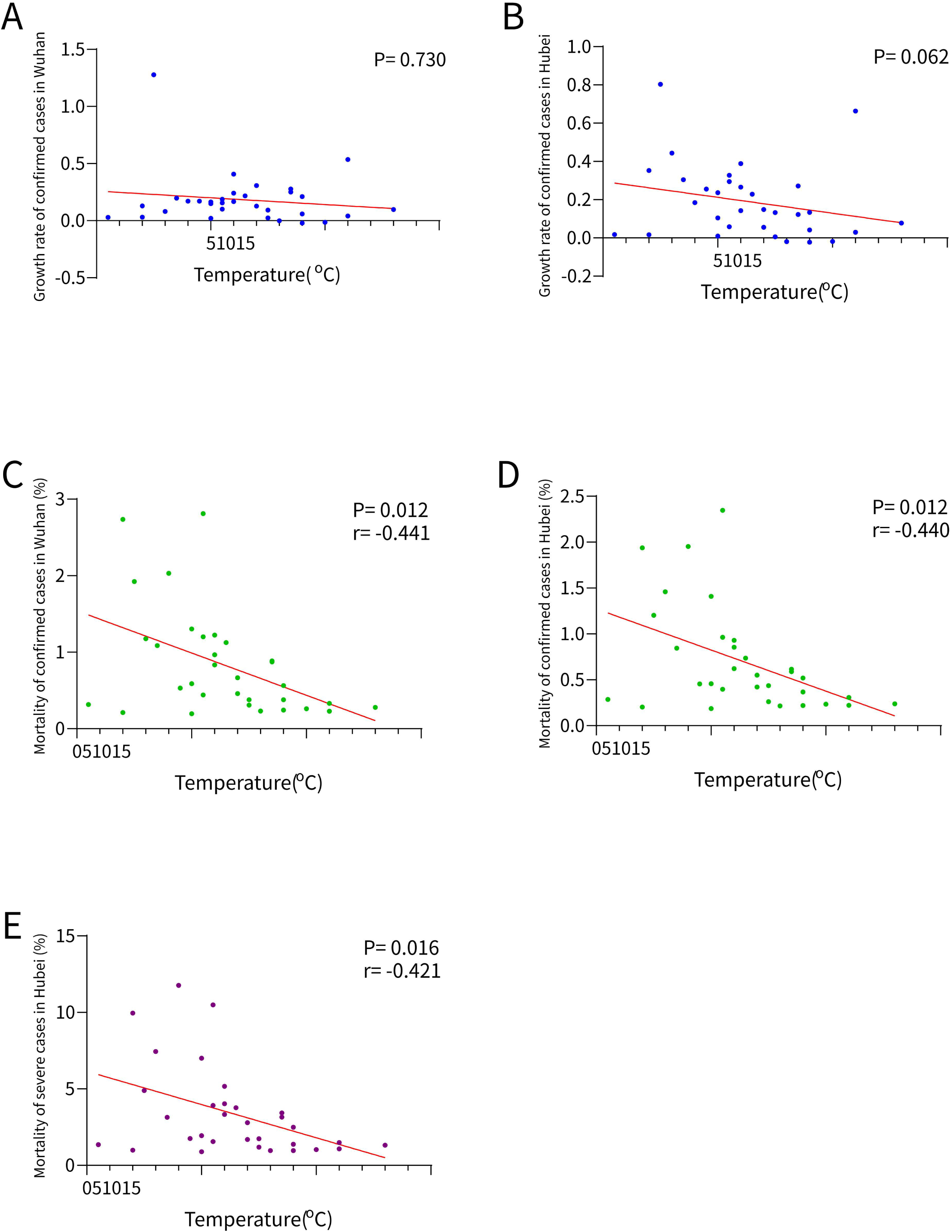
Correlation between temperature and outcomes. (**A**): correlation between air temperature and the growth rate of confirmed cases in Wuhan. (**B**): correlation between air temperature and the growth rate of confirmed cases in Hubei. (**C**): correlation between air temperature and the mortality of confirmed cases in Wuhan. (**D**): correlation between air temperature and the mortality of confirmed cases in Hubei. (**E**): correlation between air temperature and the mortality of severe cases in Hubei. Before: before the use of MSHs. After: after the use of the use of MSHs.

## Discussion

Our study found that after eight days of the use of MSHs, the mortality of COVID-19 cases in Wuhan and Hubei was significantly decreased, while as a contrast, the change in non-Hubei regions was not significant. The results preliminarily verified that these MSHs were beneficial to the survival of COVID-19 cases. Later, after another eight days, the difference of mortality between the pre and post use of MSHs was still significant in Wuhan and Hubei. However, not like the results of the first eight days, the difference in non-Hubei regions became significant. The accumulation of treatment experience of medical staff might be one of the reasons. In addition, according to the trade-off hypothesis, a pathogen must multiply within the host to ensure transmission, while simultaneously maintaining opportunities for transmission by avoiding host morbidity or death.^10^ That’s to say, coronavirus with weak virulence was more likely to spread than that with strong virulence, which might explain why the mortality in non-Hubei regions also decreased over time. However, empirical evidence remains scarce and the truth needs to be investigated.

Our study also found that the rise of AT could significantly reduce the mortality of both common and severe patients. According to a previous study, the first occurred deaths were mainly elderly people who had comorbidities or surgery history before admission.^11^ Acute or chronic cold exposure, however, could elicit bad effects on the respiratory system. Pulmonary mechanics would be compromised by bronchoconstriction, airway congestion, secretions and decreased mucociliary clearance.^12^ It has also been reported that cold exposure was coincided with hormonal changes, which might directly or indirectly alter the immune system.^13^ The above factors would worsen the underlying medical conditions of elderly people, and this might explain why a warm weather could reduce the mortality of COVID-19 cases.

When it comes to the transmissibility of coronavirus, an *in vitro* study of transmissible gastroenteritis virus and mouse hepatitis virus found that at higher AT, these coronaviruses survived for a shorter time on the surfaces of stainless steel.^14^ A case-crossover analysis performed in Saudi Arabia also found that primary Middle East Respiratory Syndrome were more likely to occur when condition was relatively cold and dry.^15^ However, our results seemed to be inconsistent with these previous studies, as no significant correlation between AT and the growth rate of confirmed cases in Wuhan was found. One of the potential reasons was that the current AT was not high enough to exert a significant impact on SARS-CoV-2. As the AT gets higher, subsequent studies are necessary to further validate our results. But considering the new cases in tropical areas such as Singapore and Malaysia,^16^ it seems that the rise of AT alone would not completely control this epidemic.

## Data Availability

1. National Health Commission of the people's Republic of China (http://www.nhc.gov.cn/)
2. China Weather Net (http://www.weather.com.cn/)

## Contributors

Yuwen Cai and Tianlun Huang made substantial contributions to conception and design, acquisition of data and analysis and interpretation of data. Xin Liu drafted the article; Gaosi Xu revised it critically for important intellectual content. All authors approved the final manuscript and agreed to be accountable for all aspects of the work.

## Transparency statement

We affirm that the manuscript is an honest, accurate, and transparent account of the study being reported; that no important aspects of the study have been omitted; and that any discrepancies from the study as originally planned have been explained.

## Data sharing statement

All data used in this study are available on the official website of National Health Commission of the people’s Republic of China (http://www.nhc.gov.cn/) and China Weather Net (http://www.weather.com.cn/).

